# Clinical and Economic Impact of Widespread Rapid Testing to Decrease SARS-CoV-2 Transmission

**DOI:** 10.1101/2021.02.06.21251270

**Authors:** A. David Paltiel, Amy Zheng, Paul E. Sax

## Abstract

**Background:** The value of frequent, rapid testing to reduce community transmission of SARS-CoV-2 is poorly understood.

**Objective:** To define performance standards and predict the clinical, epidemiological, and economic outcomes of nationwide, home-based, antigen testing.

**Design:** A simple compartmental epidemic model estimated viral transmission, clinical history, and resource use, with and without testing.

**Data Sources:** Parameter values and ranges informed by Centers for Disease Control guidance and published literature.

**Target Population:** United States population.

**Time Horizon:** 60 days.

**Perspective:** Societal.Costs include: testing, inpatient care, and lost workdays.

**Intervention:** Home-based SARS-CoV-2 antigen testing.

**Outcome Measures:** Cumulative infections and deaths, numbers isolated and/or hospitalized, and total costs.

**Results of Base-Case Analysis:** Without a testing intervention, the model anticipates 15 million infections, 125,000 deaths, and $10.4 billion in costs ($6.5 billion inpatient; $3.9 billion lost productivity) over a 60-day horizon. Weekly availability of testing may avert 4 million infections and 19,000 deaths, raising costs by $21.5 billion. Lower inpatient outlays ($5.9 billion) would partially offset additional testing expenditures ($12.0 billion) and workdays lost ($13.9 billion), yielding incremental costs per infection (death) averted of $5,400 ($1,100,000).

**Results of Sensitivity Analysis:** Outcome estimates vary widely under different behavioral assumptions and testing frequencies. However, key findings persist across all scenarios: large reductions in infections, mortality, and hospitalizations; and costs per death averted roughly an order of magnitude lower than commonly accepted willingness-to-pay values per statistical life saved ($5-17 million).

**Limitations:** Analysis restricted to at-home testing and limited by uncertainties about test performance.

**Conclusion:** High-frequency home testing for SARS-CoV-2 using an inexpensive, imperfect test could contribute to pandemic control at justifiable cost and warrants consideration as part of a national containment strategy.

**Primary Funding Sources:** Dr. Paltiel was supported by grant R37DA015612 from the National Institute on Drug Abuse of the National Institutes of Health.

Dr. Sax was supported by grant R01AI042006 from the National Institute of Allergy and Infectious Diseases of the National Institutes of Health.

## INTRODUCTION

Previous research identified social distancing (masks, de-densification, lockdowns), large-scale diagnostic testing, and vaccination as essential elements of a coordinated plan to contain the COVID-19 pandemic. One strategy receiving less formal attention is the high-frequency use of low-cost, rapid, home-based antigen testing for SARS-CoV-2 with self-enforced isolation for those who obtain a positive result.^1-5^ Advocates point to the potential advantages: prevention by isolation; focusing on “infectiousness” rather than “infection”; and reducing strain on laboratories that must conduct viral polymerase chain reaction (PCR)-based diagnostic testing.^6-8^ Critics note that such an approach risks poor uptake and adherence, frequent false negatives leading to unfounded reassurance, and frequent false positives resulting in needless isolation and lost work productivity.^9^

We seek to formalize this discussion by capturing both the promise and the concerns in the structure of a mathematical model. We aim to identify the circumstances that would have to prevail – about the accuracy of antigen testing, about the cost of both initial and confirmatory tests, about the lost workdays arising from isolation (both true and false positives) and disease, and about individual behavior – for such an intervention to be of clinical, economic, epidemiologic, and policy interest.

## METHODS

### Study Design

We adapted a simple compartmental epidemic model (**Appendix Figure A1**) to capture the essential elements of a population-wide, home-based testing program to detect, isolate, and contain contagious SARS-CoV-2. Features we sought to portray included: the epidemiology of infection; the natural history of COVID-19 illness; the behavioral response to test availability, test results, and isolation; and the financial consequences of testing, hospitalization, and lost workdays arising from illness, isolation, and (potentially incorrect) test findings. A spreadsheet implementation of the model permitted us to vary critical input data parameters and to examine how different test performance attributes (e.g., frequency, sensitivity, specificity, cost), different behavioral responses (e.g., acceptance of antigen testing, willingness to self-isolate, propensity to abandon isolation), and different epidemiological scenarios would translate into both health outcomes (e.g., tests administered, true/false positives, new infections, person-days requiring isolation, hospitalizations, and deaths) and economic performance (e.g., testing costs, inpatient costs, lost productivity, and cost-effectiveness). Given the rapid spread of infection and the speed of new advances, we adopted a short, 60-day planning horizon.

Input data (**Appendix Table A1**) were obtained from published sources, adhering whenever possible to planning scenarios and data guidance for modelers from the Centers for Disease Control and Prevention (CDC) and the Office of the Assistant Secretary for Preparedness and Response (ASPR).^10-20^ Because our aim was to identify the circumstances under which widespread, rapid home testing might warrant inclusion as part of a national containment strategy, we deliberately tipped the scales to portray the intervention in a less favorable light, choosing false-positive and false-negative rates at the upper end of the plausible range, inflating costs, and exaggerating levels of non-compliance with recommended protocols. To provide context for our estimates of the incremental cost per death averted, we followed the guidance of the Office of the Assistant Secretary for Planning and Evaluation and applied the value of a statistical life (VSL), a benchmark of the societal willingness to pay for reductions in mortality risks.^10^ Here again, we erred on the side of conservatism, adopting the lower bound value ($5.3 million) from the recommended range (central estimate = $11.5 million; upper bound = $17.3 million).

### Compartmental Model

We made two notable changes to the traditional “susceptible-exposed-infected-removed” (or “SEIR”) compartmental modeling framework (**Appendix Figure A1**). First, we separated the single “infected” compartment into four sub-compartments: “asymptomatic”, “mild/moderate”, “severe”, and “critical”. This permitted us to capture more fully the natural history, epidemiology, and resource use associated with progressive COVID-19. Second, we distinguished between epidemiologically “active” and “removed” individuals. In “active” compartments, we assumed that individuals interact in ways that permit infectious contact and transmission of SARS-CoV-2; in “removed” compartments, no transmission was possible. We provided two pathways into the “removed” compartments. Individuals with “severe” and “critical” infection were removed by virtue of their advanced COVID-19 symptoms and hospitalization. (A user-defined variable also permitted some fraction of individuals with “mild/moderate” symptoms to self-isolate.) As described in greater detail in the next section, testing offered a second avenue to removal.

We considered different background epidemic severities. We assumed baseline effective reproduction number R_t_ = 1.3 and explored values ranging from 0.9 to 2.1 in sensitivity analysis. As detailed in the **Appendix**, we assumed that 60% of infections would produce symptoms, that 10% of those with symptoms would advance to severe disease, and that 5% of those with severe disease would advance to critical illness. Base occupancy times for persons progressing to more advanced illness were 3, 10, 6, and 4 days in the “exposed”, “asymptomatic”, “mild/moderate”, and “severe” states, respectively. We used mortalities of 1%, 5%, and 15% from the “mild/moderate”, “severe”, and “critical” states.

### Performance of Testing

Regular opportunities to be tested for SARS-CoV-2 contagion were offered to persons in the active “uninfected”, “exposed”, “asymptomatic”, and “mild/moderate” compartments. We reviewed the data on the performance of antigen testing and found sensitivity estimates ranging from a low of 41.2% to a high of 100% and specificities from 97% to 99.9%.^7, 21-27^ We then focused our estimates on cases when testing was accompanied by either a low cycle threshold (high titers of virus) or the ability to isolate replication-competent virus, as these cases represented people in the most transmissible stage of COVID-19 and excluded those in the recovery phase who might still have positive PCRs.^13, 28-29^ While some studies described antigen sensitivity as exceeding 90% under these circumstances, we deliberately lowered our base case sensitivity assumption to 80% to reflect less-than-optimal testing characteristics in the home setting. We did not consider how repeat testing might increase this sensitivity. Similar motivations led us to adopt a lower-bound value of 95% for test specificity, exploring even lower values (90%) in sensitivity analysis.^7^ We examined weekly testing in the base case but considered frequencies ranging from daily to once every 15 days in sensitivity analysis.

To capture both the costs and the potential delays of confirmatory testing, we assumed that the shipment of testing kits to households would include a swab for obtaining a PCR test, to be self-collected at home and sent to a central laboratory in the event of a positive rapid test result. Individuals whose initially false-positive result had led them to adhere to recommended isolation protocols were assumed to return to the active population after a 3-day delay.^30^ (This assumption ignored the small possibility of repeatedly false-positive confirmatory test findings.)

Finally, we assumed that, in the absence of disease progression, successful isolation of infected persons for 10 days would lead to recovery and non-infectious return to the “active” population.

### Behavioral response

To account for concerns about individual willingness to adhere to testing and isolation protocols, we adopted a highly pessimistic view of the behavioral response to the testing intervention. In the base case, we assumed that: a) only 50% of individuals would elect to make use of the test kits provided to them; b) only 50% of individuals receiving a positive test finding would respond by isolating themselves as instructed; and c) even among those who did initially isolate, 20% each day would abandon isolation and return to the “active” population against recommended guidance. We further assumed that only 50% of persons exhibiting moderate symptoms of COVID-19 would elect to self-isolate, in the absence of positive test finding. Here again, we assumed that even among symptomatic individuals who did initially elect to isolate, 20% would abandon isolation each day.

Recognizing the uncertainties surrounding these assumptions, we examined a spectrum of alternative behavioral response scenarios – including an even-more-pessimistic “worst case” – in sensitivity analysis (**Table 1**).

**Table 1.** Behavioral scenarios

|  | Testing Behavior Scenario |  |  |
| --- | --- | --- | --- |
|  | Best Case | Base Case | Worst Case |
| % of individuals who make use of tests offered | 80 | 50 | 25 |
| % of individuals who self-isolate after a positive test result | 75 | 50 | 25 |
| Daily % of individuals who abandon isolation | 5 | 20 | 33 |

### Economic outcomes

We assigned 3 categories of cost: testing, inpatient, and lost productivity. Here again, we chose values that would deliberately bias the analysis against the intervention: exaggerating the costs of testing and lost workdays, and under-stating the costs of hospitalization. Testing costs included both the initial testing kit ($5, range $1-$10) and the confirmatory test (base $20, range $5-$50). The current price of a single over-the-counter antigen test for SARS-CoV-2 is roughly $25 or more, a figure that includes a substantial markup for both the manufacturer and the retailer and that offers no quantity discount.^17^ Experience with other infectious diseases (malaria, for example) suggests that high-volume, paper-strip-based antigen tests can be obtained at wholesale prices as low as $0.20 per unit for government-based purchase orders of the magnitude being considered here.^18^ The initial test costs were assigned on an “intent-to-treat” basis. In other words, mailing test kits to someone’s home incurred the cost, whether or not the individual chose to use the test. Inpatient costs were assigned per day with severe illness ($1,000) or critical illness ($2,500). Every day spent in the hospital or in isolation – regardless of whether isolation was the result of a true- or a false-positive finding – was treated as a day of lost productivity, which we assigned a cost of $180, based on the daily *per capita* gross domestic product.^19,20^

## RESULTS

### Base case

In the absence of a testing intervention, the model anticipates 15 million infections, 125,000 deaths, and $10.4 billion in costs ($6.5 billion inpatient; $3.9 billion lost productivity) over a 60-day horizon (**Table 2**). Weekly home testing under base case assumptions could reduce infections to 11 million and deaths to 106,000. Lower inpatient costs ($5.9 billion) would partially offset additional outlays for testing ($12.0 billion) and greater lost workdays ($13.9 billion). Although testing and isolation would increase the total number of workdays lost (from 21.4 million to 77.4 million days), individuals could expect to be isolated 0.17 days (1.1 days under the most pessimistic possible data assumptions) owing to false-positive findings. Compared to the status quo, the testing intervention would produce a cost per infection averted of $5,400 and a cost per death averted of $1.1 million. Applying the lowest-available estimate from the recommended range of VSL benchmark values ($5.3 million), this suggests that the intervention would be an exceptionally good value.

**Table 2.**
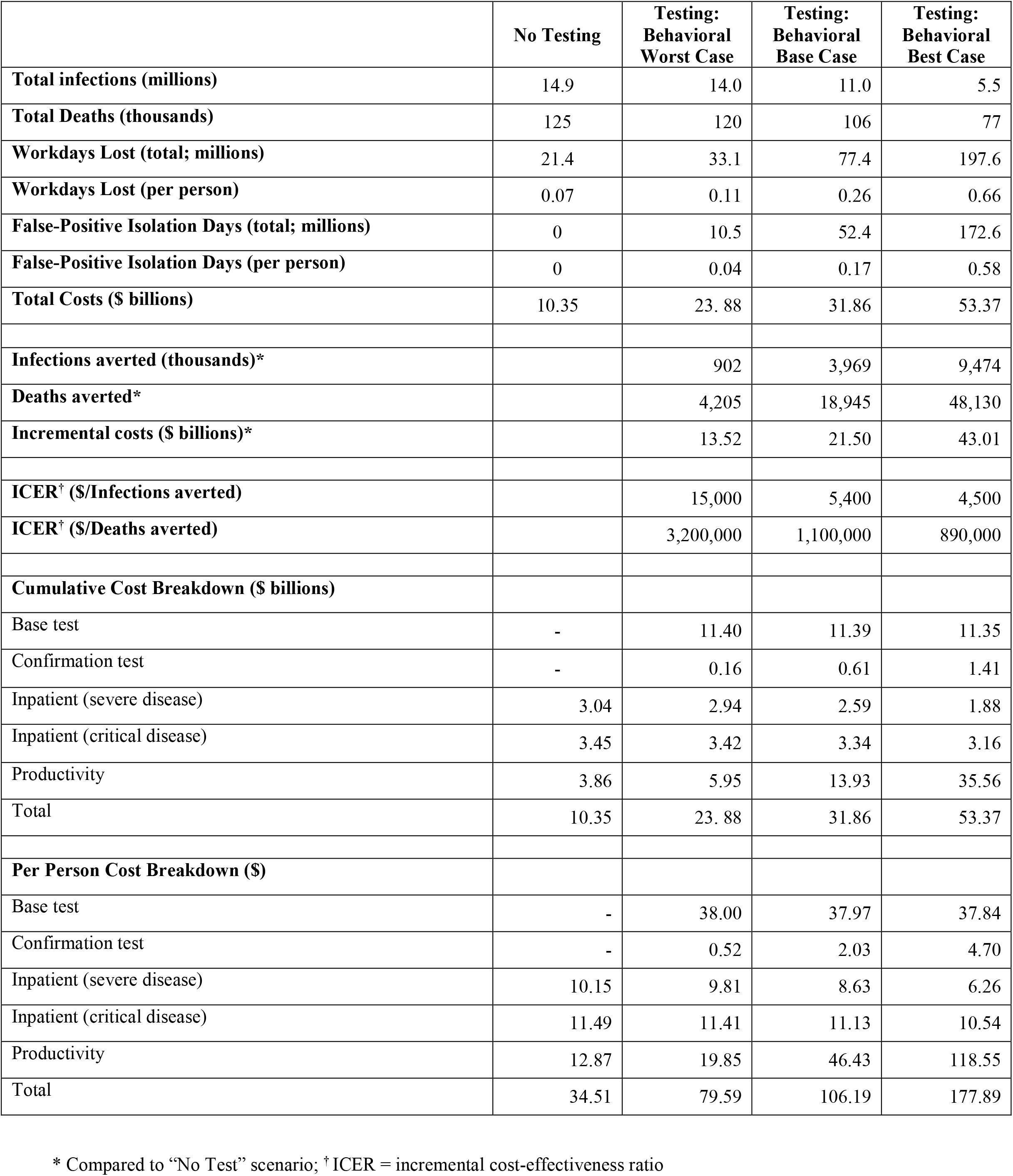
Clinical and Economic Outcomes Under Alternative Testing and Behavior Assumptions

### Sensitivity to Behavioral Factors

Test acceptance and adherence to isolation protocols would greatly influence the magnitude of all estimated outcomes (**Figure 1 and Table 2**). Under Best Case behavioral assumptions, the testing intervention alone could begin to contain the epidemic; under less favorable behavioral scenarios, the impact of testing on the spread of infection would be less pronounced. However, even under the Worst Case behavioral scenario (25% participation; 25% isolation of positives; 33% rate of daily abandonment), more than 900,000 infections could be prevented over 60 days.

**Figure 1.**
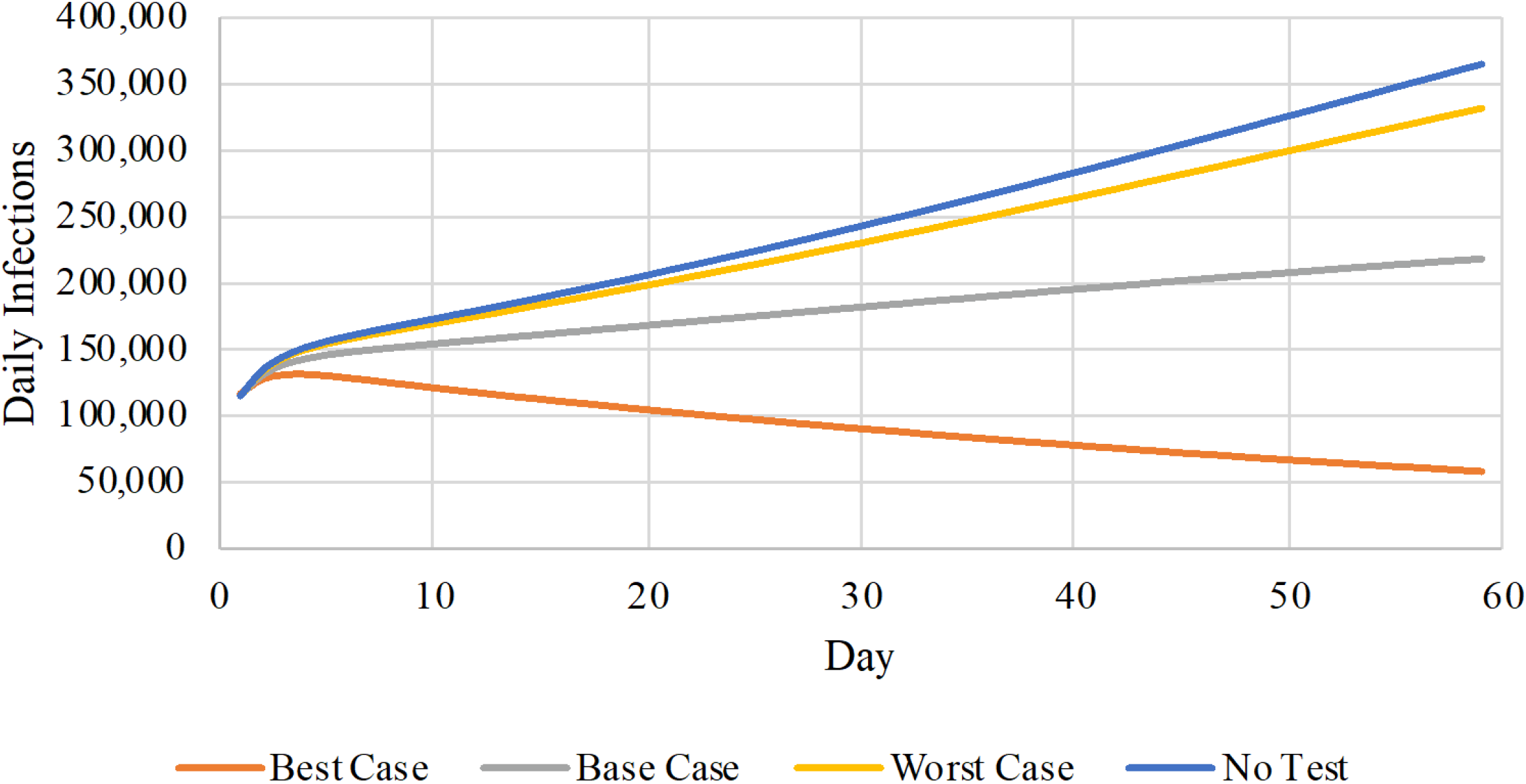
Daily infections as a function of behavioral scenarios. This figure reports the daily number of infections (vertical axis) under three behavioral scenarios with home-based testing and no home-based testing over a 60-day horizon (horizontal axis). The colored lines denote different testing and behavioral assumptions: no testing (blue); best case (orange); base case (gray); and worst case (yellow).

Costs too would vary widely depending on behavioral assumptions (**Table 2**). Greater adherence to program protocols would invariably produce greater overall costs, comprised of higher costs of testing (initial and confirmatory) and higher costs of lost workdays, partially offset by lower costs of inpatient care. But cost-effectiveness ratios (e.g., costs per infection and per death averted, compared to No Testing) would behave more stably. Even under the most pessimistic behavioral assumptions, the incremental cost per death averted ($3.2 million) would remain below the most stringent recommended benchmark VSL ($5.3 million).

### Sensitivity to Test Frequency

Figure 2. illustrates the critical role played by test frequency. A test that elicits a poor behavioral response will still prevent a large number of infections, if offered with sufficient frequency. Even under Worst Case behavioral assumptions, for example, cumulative infections could be cut more than 33% via the daily offer of testing.

### Sensitivity to R_t_

Under all scenarios and testing assumptions, greater epidemic severity (as measured by the reproductive number, R_t_) would produce more infections, more deaths, and higher costs. It would also engender more favorable results for the testing intervention, as measured by infections averted, by deaths averted, and by cost-effectiveness ratios (**Table 3**).

**Table 3.** Sensitivity of key outcomes to epidemic severity

| | **Total Cost (\$)** | **Total infections** | **Total deaths** | **Incremental cost (\$)\*** | **Infections averted\*** | **Deaths averted\*** | **ICER<sup>†</sup> (\$/Infections averted)** | **ICER<sup>†</sup> (\$/Deaths averted)** |
| --- | --- | --- | --- | --- | --- | --- | --- | --- |
| **R<sub>t</sub> = 0.9** |  |  |  |  |  |  |  |  |
| No Test | 6,423,408,708 | 3,856,513 | 65,380 |  |  |  |  |  |
| Base Case | 28,265,635,826 | 3,084,105 | 60,406 | 21,842,227,118 | 772,408 | 4,974 | 28,278 | 4,390,952 |
| **R<sub>t</sub> = 1.3** |  |  |  |  |  |  |  |  |
| No Test | 10,351,589,938 | 14,930,587 | 124,637 |  |  |  |  |  |
| Base Case | 31,856,176,855 | 10,961,643 | 105,692 | 21,504,586,917 | 3,968,943 | 18,945 | 5,418 | 1,135,079 |
| **R<sub>t</sub> = 1.7** |  |  |  |  |  |  |  |  |
| No Test | 21,094,569,851 | 53,002,937 | 286,342 |  |  |  |  |  |
| Base Case | 41,662,061,788 | 38,203,510 | 227,026 | 20,567,491,937 | 14,799,426 | 59,317 | 1,390 | 346,740 |
| **R<sub>t</sub> = 2.1** |  |  |  |  |  |  |  |  |
| No Test | 44,402,386,231 | 131,798,989 | 638,154 |  |  |  |  |  |
| Base Case | 64,250,192,494 | 103,668,349 | 504,479 | 19,847,806,264 | 28,130,639 | 133,675 | 706 | 148,478 |

\* Compared to “No Test” scenario
<sup>†</sup> ICER = incremental cost-effectiveness ratio

**Figure 2.**
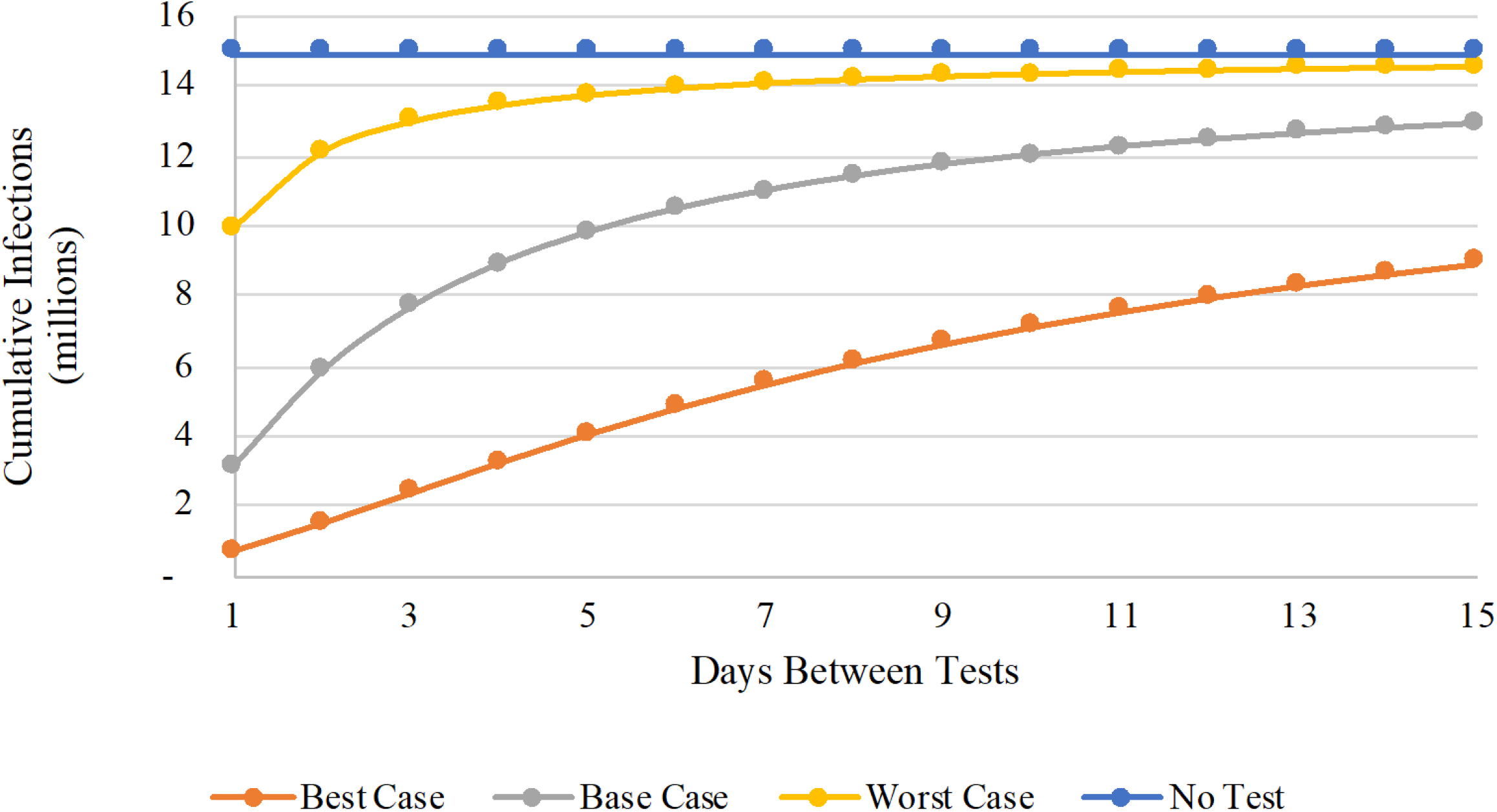
Cumulative infections as a function of testing frequency. In this figure, the number of cumulative infections (vertical axis, in millions) is reported for a range of home-based testing frequencies (horizontal axis, ranging from 1 to 15 days between tests). The colored lines denote different testing and behavioral assumptions: no testing (blue); best case (orange); base case (gray); and worst case (yellow).

### Sensitivity to the costs of testing

With initial and confirmatory test costs set to their base case values ($5 and $20), the intervention had an incremental cost-effectiveness ratio of $1.1 million/death averted. Setting testing costs to the lowest ($1, $10) and highest ($10, $50) values in our estimated range yielded ICERs per death averted of $630,000 and $1.8 million, respectively.

### Size and composition of isolated population

Regardless of the testing protocol, large numbers of people will be required to isolate. In the absence of testing, we expect to observe 21 million person-days spent in isolation, all of them attributable to symptoms and hospitalization (**Table 2**). Notably, this means that the average individual can expect to lose 0.07 workdays to isolation. Under base case assumptions, testing will increase total days spent in isolation to 77 million (or 0.26 days per person). However, only 22% of those days will be attributable to hospitalization, the remainder being the result of testing (11% true positives; 68% false positives). Under the most pessimistic possible assumptions (i.e., best-case adherence to isolation protocols coupled with worst-case test performance), the average member of the population can expect to spend 1.1 days in unnecessary isolation (i.e., for a false positive) over the course of the 60-day horizon.

## DISCUSSION

This model-based analysis finds that a nationwide program of frequent, antigen-based home testing and self-isolation could greatly reduce total infections and mortality at a justifiable cost. We arrive at this conclusion using the methods of cost-effectiveness analysis and assumptions that were chosen with the deliberate intent of portraying all aspects of the intervention – the performance of antigen testing; the behavioral response of individuals to testing and isolation protocols; and the societal willingness to pay to avert untimely deaths – in an unfavorable light. Use of more middle-of-the-road data assumptions would only serve to strengthen our policy conclusion. Our bottom line message: do not let the perfect be the enemy of the good; even a highly imperfect home-based testing program could confer enormous benefit.

With an analysis that uses the entire US population as its target, the numerical results reported here are staggering in their magnitude, sensitive to small changes in the input values, and difficult to digest. Although we have attempted to address this by reporting values on a *per capita* basis and by conducting extensive sensitivity analysis, we nevertheless urge the reader to focus less on our numerical point estimates and more on the remarkable robustness of our qualitative, policy finding: namely, that a nationwide rollout of frequent, home-based testing and self-isolation is justified on both epidemiological and economic grounds.

In addition to the many limitation surrounding any model-based evaluation, our analysis does not account for the potential benefits of regular, inexpensive, rapid testing in other, more targeted settings. Schools, factories, air travel, concerts, recreational sports, large family celebrations, and places of worship might choose to make such testing a pre-requisite of participation. Since a small number of people with COVID-19 account for a high proportion of secondary cases, such a strategy would remove some of the most contagious individuals from crowded settings, eliminating case clusters and preventing super-spreader events.^31^

Some observers have questioned the ability of frequent, rapid, antigen testing to reduce transmission, raising several concerns.^9^ These include the lower sensitivity of antigen testing compared to PCR-testing (raising the risk that infectious people will remain in public on the erroneous belief that they are not infectious), the high number of false-positive tests leading to unnecessary isolation, poor adherence to the recommended testing and isolation recommendations, and the massive expense if testing is broadly applied. We acknowledge these concerns and, wherever possible, we have tried to give them voice by adopting modeling assumptions and input data values that tip the scales against nationwide antigen-based home testing.

The strategy of frequent rapid testing to reduce SARS-CoV-2 transmission and decrease COVD-19 cases began generating widespread attention in the popular press and on social media many months ago.^32,33^ Implementation on a population-based level, however, has been limited to date. A program of massive, rapid, antigen testing in Slovakia on consecutive weekends appears to have contributed to a reduction in COVID-19 cases beyond what would have been expected through standard infection control measures.^34^ Initiation of a community-based testing pilot in

Liverpool was associated with a decline in cases, but it is not clear if this was the result of testing or increased infection-control measures and other restrictions.^35^ Current obstacles to broader use of these tests in the United States include the requirement for a doctor’s order for certain tests, limited availability, and continued high prices.

Our findings confirm those of other investigators that home-based rapid testing can have a dramatic effect in reducing transmission and COVID-19 mortality at a justifiable cost,^1,32^ with testing frequency a key component to why this broad testing strategy is so effective. Factors currently in play that could worsen the pandemic – a slow start to roll out of widespread vaccination, “pandemic fatigue” regarding social distancing and mask wearing, and emergence of variants that are more easily transmissible – only underscore the importance of frequent rapid testing as a strategy to reduce the number of new cases.

## Data Availability

This study makes secondary use of previously published data. All sources are listed in the manuscript tables and references.

## Technical Appendix

### Model Overview

We developed a dynamic, compartmental model using a modified “susceptible-exposed-infected-recovered” (or SEIR) framework. The model portrays the epidemiology and natural history of infection in a homogeneous population of at-risk individuals as a sequence of transitions, governed by difference equations, between different health states (or “compartments”). The model diagram (**Appendix Figure 1**, below) illustrates the modifications we made to the basic SEIR framework:

- Division of the “Infected” state into four distinct sub-compartments, to capture the increasing severity and hospital resource use associated with more advanced COVID-19 disease: “Asymptomatic,” “Mild/Moderate” (outpatient), “Severe” (hospitalized) and “Critical” (hospitalized in an intensive care unit [ICU]). Transitions representing disease progression are denoted in the diagram with green arrows.
- Introduction of a parallel set of states to distinguish between epidemiologically “active” and “removed” individuals. In “active” compartments (on the right side of the diagram), we assumed that individuals interact in ways that permit infectious contact and transmission of SARS-CoV-2; in “removed” compartments (left side), no transmission was possible.

This gave rise to a total of 13 model states

- U: Uninfected, active
- E: Exposed, asymptomatic, active
- A: Infected, asymptomatic, active
- M: Infected, mild-moderate illness, outpatient, active
- S: Infected, severe illness, inpatient, removed based on hospitalization
- C: Infected, critical illness, inpatient, removed based on hospitalization
- U_FP:_ Uninfected, removed base on false positive test
- E_FP_: Exposed, asymptomatic, removed base on false positive test
- A_TP_: Infected, asymptomatic, removed base on true positive test
- M_Obs_: Infected, mild-moderate illness, outpatient, removed based on observed symptoms
- M_TP_: Infected, mild-moderate illness, outpatient, removed base on true positive test
- R: Recovered, non-infectious, active
- D: Dead, removed

### Removal

We provided two pathways into the “removed” compartments. Individuals with “severe” and “critical” infection were immediately removed by virtue of their advanced COVID-19 symptoms and hospitalization. (We assumed that proper hospital infection control policies would prevent any nosocomial transmission; thus, persons in states S and C do not transmit infection in the model.) We further assumed that that some fraction of individuals with “moderate” symptoms would also elect to self-isolate and therefore enter the “removed” states. (We held this fraction constant at 50% in the present analysis.)

Testing offered a second avenue to removal. Regular opportunities to be tested for SARS-CoV-2 contagion were offered to persons in the active “uninfected”, “exposed”, “asymptomatic”, and “mild/moderate” compartments. All transitions into the removed compartments arising from positive tests are denoted in the diagram with orange arrows.

- Uninfected individuals receiving a false positive test result moved from the uninfected state (U) to the uninfected/false positive state (U_FP_).
- Similarly, individuals exposed to infection (and not yet identifiable as infected via testing) receiving a false positive test result moved from the exposed state (E) to the exposed/false positive state (E_FP_).
- Asymptomatic individuals receiving a (true) positive test result moved from state (A) to state (A_TP_).
- Individuals with Mild/Moderate illness (state M) who elected not to self-isolate based upon their symptoms could nonetheless transition to state M_TP_ after receiving a (true) positive test result.

Importantly, a positive test result alone was not enough to trigger transition to a removed state. To account for concerns about individual willingness to adhere to testing and isolation protocols, we adopted a highly pessimistic view of the behavioral response to the testing intervention. We assumed that: a) only some fraction (base case = 50%) of individuals would elect to make use of the test kits provided to them; b) only some fraction (base case = 50%) of individuals receiving a positive test finding would respond by isolating themselves as instructed.

### Return to active states

We provided three pathways from the “removed” compartments back into the “active” population:

- “Abandonment”. We assumed that among individuals who initially isolated in response to a positive result, some fraction (base case = 20%) would abandon isolation each day and return to the “active” population, against recommended guidance. Because of the short incubation time for COVID-19, we assumed that abandonment of state E_FP_ would result in a return to state A. For all other “removed” states, abandonment resulted in a return to the analogous “active” state (i.e., from U_FP_ to U; from A_TP_ to A; and from M_Obs_ and M_TP_ to M). All transitions arising from abandonment of isolation are denoted in the diagram with purple arrows.
- Confirmatory testing. We assumed that the shipment of testing kits to households would include a swab for obtaining a PCR test to be self-collected at home and sent to a central laboratory in the case of a positive rapid test result. Individuals whose initially false-positive result had led them to adhere to recommended isolation protocols (states U_FP_ and E_FP_) were assumed to return to the active population after an average 3-day delay. Transitions arising from confirmatory testing are denoted in the diagram with yellow arrows.
- Recovery. We assumed that, in the absence of disease progression, successful isolation of infected persons in states A_TP,_ M_Obs_ and M_TP_ for an average of 10 days would lead to recovery and non-infectious return to the “active” population. (Note: in the absence of disease progression, transition to the Recovered state was also possible for persons in active states A and M, after an average of 10 days.) Persons in the recovered state R were assumed to remain active in the transmission pool but are neither able to transmit nor to be infected a second time. All transitions arising from recovery are denoted in the diagram by dotted blue arrows.

### Death

COVID-related mortality was possible from any of states M, M_Obs_, M_TP,_ S, and C. All transitions arising from death are denoted in the diagram by dotted red arrows.

### Parameters

β_i_: rate at which infected individuals in state i contact susceptibles and infect them. This applies to states A and M.

p_i_: rate of progression from disease state i to the next stage of disease. This applies to states E, A, M, S, A_TP_, M_Obs_, and M_TP_.

r_i_: rate at which individuals in state i recover from disease. This applies to states A, M, S, C, A_TP_, M_Obs_, and M_TP_.

m_i_: mortality rate for individuals in state i. This applies to states M, S, C, M_Obs_, and M_TP_.

v_i_: testing rate for individuals in state i. This applies to states U, E, A, and M. This rate is a multiplicative composite of the base frequency of test offers and the likelihood that an individual in state i will accept a test offer. (We assumed the same value for individuals in all states.)

ℓ_i_: isolation probability for individuals in state i who receive a positive test result. This applies to states U, E, A, and M. We also apply this probability to persons who develop Mild/Moderate symptoms. (We assumed the same value for individuals in all states.)

λ_i_: isolation abandonment rate for individuals in state i. This applies to states U_FP_, E_FP_, A_TP_, M_Obs_, and M_TP_. (We assumed the same value for individuals in all states.)

κ_i_: false-positive confirmatory test return rate for individuals in state i. This applies to states U_FP_ and E_FP_. (We assumed the same value for individuals in all states.)

Se / Sp: sensitivity and specificity of the home antigen test The model uses a cycle time of 1 day. All rates are calculated per daily cycle.

### Governing equations

For ease of computation, we define

- **Weighted force of infection at time t**

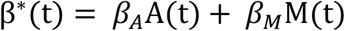 **Active population at time t**

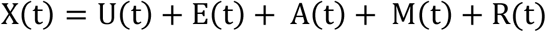

The equations governing transitions from one state to the next are:

- **Uninfected (t+1) = Uninfected (t) – New Exposures to E – Isolating False Positives + Returning False Positives (confirmatory test and balking)**

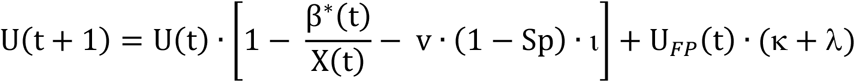

- **Exposed (t+1) = Exposed (t) – New Infections to A – Isolating False Positives + New Exposures from U**

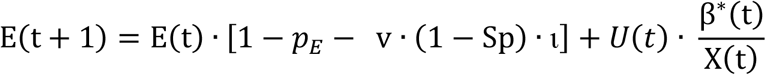

- **Asymptomatic (t+1) = Asymptomatic (t) – Progressions to M – Isolating True Positives – Recoveries + New Infections from E + Returning False Positives from E**_**FP**_ **(confirmatory test and balking) + Balking True Positives from A**_**TP**_

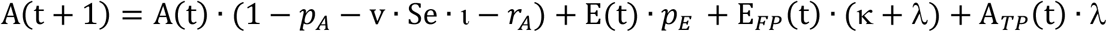

- **Mild/Moderate (t+1) = Mild/Moderate (t) – Progressions to S – Recoveries – Deaths – Isolating True Positives + Progressions from A + Balking True Positives from M**_**Obs**_ **+ Balking True Positives from M**_**TP**_

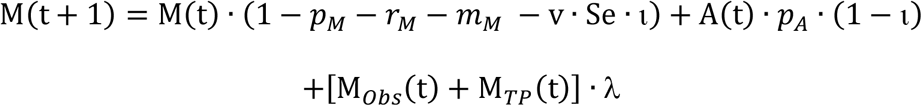

Only fraction (1 - ι) of individuals progressing to Mild/Moderate symptoms from A will enter state M. The other fraction (ι) will directly self-isolate to state M_Obs_based on their observed symptoms.

- **Severe (t+1) = Severe (t) – Progressions to C – Recoveries – Deaths + Progressions from M, M**_**Obs**_, **and M**_**TP**_

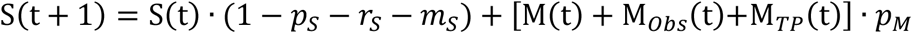
- **Critical (t+1) = Critical (t) – Recoveries – Deaths + Progressions from S**

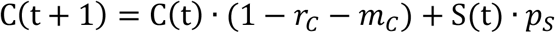
- **Uninfected False Positive (t+1) = Uninfected False Positive (t) – Returns to U (confirmatory test and balking) + Isolating False Positives**

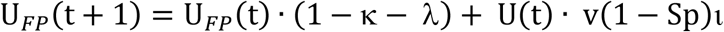
- **Exposed False Positive (t+1) = Exposed False Positive (t) – Returns to A (confirmatory test and balking) + Isolating False Positives**

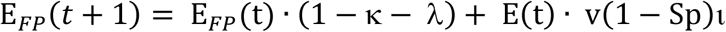
- **Asymptomatic True Positive (t+1) = Asymptomatic True Positive (t)– Progressions to M**_**TP**_ **– Recoveries – Balking returns to A + Isolating True Positives**

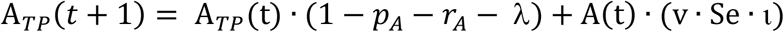
- **Mild/Moderate Observed (t+1) = Mild/Moderate Observed (t) – Progressions to S – Recoveries – Deaths – Balking returns to M + Isolating based on symptoms from A**

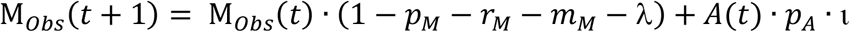

Fraction ι of individuals progressing to Mild/Moderate symptoms from A will directly self-isolate to state M_Obs_ based on their observed symptoms.

- **Mild/Moderate True Positive (t+1) = Mild/Moderate True Positive (t) – Progressions to S – Recoveries – Deaths – Balking returns to M + Progressions from A**_**TP**_ **+ Isolating True Positives from M**

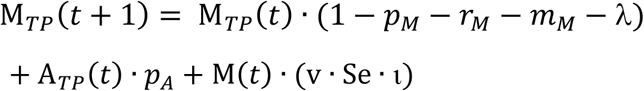
- **Recovered (t+1) = Recovered (t) + New Recoveries**

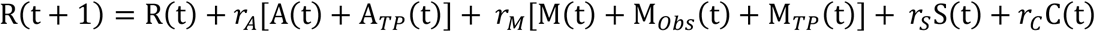
- **Deaths (t+1) = Deaths (t) + New Deaths**

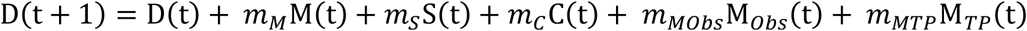
- **Total Population at time t =**

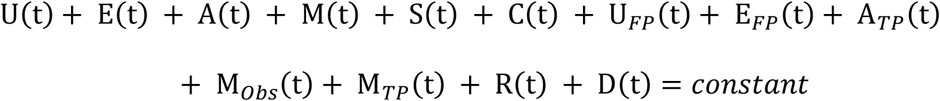

### Initial conditions

We considered the US population (N = 300 million) with the following initial distribution

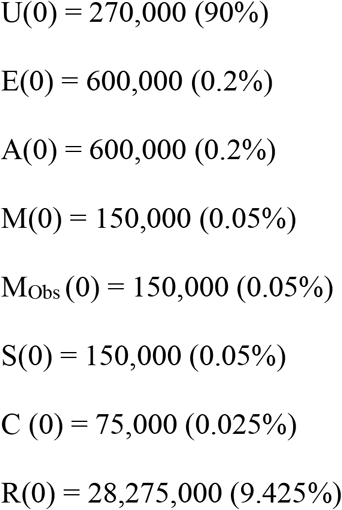

All other state compartments are empty at time 0.

### Estimating disease progression, recovery, and mortality rates

In this section, we describe how the parameters used in the governing equations are derived. Unless otherwise noted as an additional assumption, all input values contained in this section are listed and referenced in **Appendix Table 1**.

- **From Exposed state, E** Individuals spend an average of 3 days in the Exposed state before progressing to Asymptomatic infection. This implies p_E_ = (3 days)^-1^ = 0.333.
- **From Asymptomatic state, A** Assume that 40% of asymptomatic individuals recover without progressing to Mild/Moderate disease. For the 60% who do progress to Mild/Moderate disease, the average time spent in the Asymptomatic state is 10 days. This permits us to solve for both progression rate pA = (10 days)^-1^ = 0.1 and recovery rate rA = (0.4/(1-0.4))*pA = 0.067
- **From Mild-Moderate state, M** Assume that individuals spend an average of 6 days in state M. This gives us (pM + mM + rM)^-1^ = 6. Further assume that 10% of persons who enter Mild/Moderate illness will eventually progress to Severe illness. This suggests that pM / (pM + mM + rM) = 0.1 and, hence, pM = 0.1/6 = 0.0167. Finally, assume that 1% of all persons who arrive in state M will die while in that state. This means that mM/(pM + mM + rM) = 1% and, since we know that (pM + mM + rM)^-1^ = 6, then mM = 0.01/6 = 0.00167. This also permits us to solve for rM: since rM/(pM + mM + rM) = (1 – 10% - 1%) = 89%, and, since we know that (pM + mM + rM)^-1^ = 6, then rM = 0.89/6 = 0.1483.
- **From Severe state, S** Individuals spend an average of 4 days in state S. This gives us (pS + mS + rS)^-1^ = 4. Further assume that 5% of all persons who arrive in state S will die in that state. This means that mS/(pS + mS + rS) = 0.05 and hence mS = 0.05/4 = 0.0125. Now assume that 5% of persons who enter Severe illness will progress to Critical illness. This suggests that ps/(pS + mS + rS) = 0.05, implying that ps = .05/4 = 0.0125. We can now solve for the proportion of persons in state S who recover: rs/(pS + mS + rS) = (1 – 5% - 5%) = 90% and since (pS + mS + rS)^-1^ = 4, then rS = 0.9/4 = 0.225.
- **From Critical state, C** The average time spent in state C is 14 days. This gives us (mC + rC)^-1^ = 14. Further, 15% of all persons who arrive in state C will die; the other 85% will recover. This means that mC/(mC + rC) = 0.15 and hence mC= 0.15/14 = 0.0107 while rC/(mC + rC) = 0.85 and hence rC= 0.85/14 = 0.0607.

### Estimating transmission parameters

The basic reproduction number (R_t_) measures the transmission potential of an infectious agent. In the absence of a vaccine, the basic reproduction number associated with this model is given by

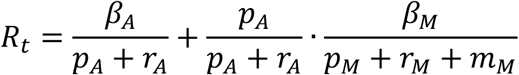

We have already derived parameters pA, rA, pM, rM, and mM, above. We assumed that asymptomatic persons were five times more likely to transmit infection than persons with mild/moderate infection and hence β_A_ = 5β_M_. We then solved for the values of β_A_ and β_M_ for a range of different reproduction numbers. In particular, for R_t_ = {0.9, 1.3, 1.8. 2.1}, we obtained β_A_ = {0.14, 0.2, 0.28, 0.33} and β_M_ = {0.028, 0.041, 0.057, 0.066}.

**Appendix Figure 1.**
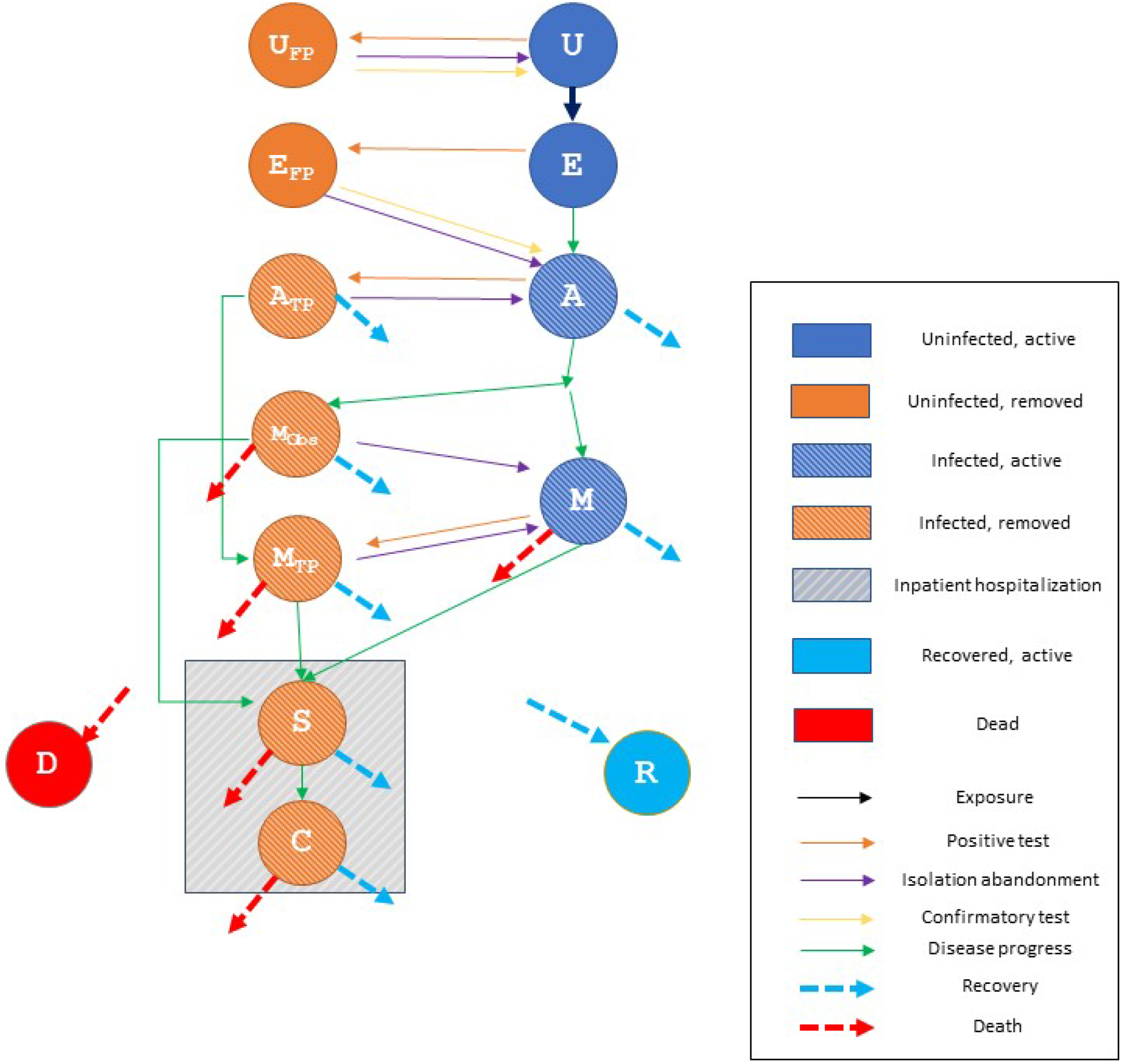
Model Structure

**Appendix Table A1.** Model parameters and inputs

| **Model inputs** |  | **References** |
| --- | --- | --- |
| **Initial population size (n)** | 300,000,000 | Assumption |
| **Compartments** | **Initial population (%)** |  |
| Non-infected, susceptible | 90.00 | Assumption |
| Exposed | 0.20 | Assumption |
| Infected, asymptomatic | 0.20 | Assumption |
| Infected, symptomatic | 0.05 | Assumption |
| Infected, symptomatic, in isolation | 0.05 | Assumption |
| Infected, hospitalized | 0.05 | Assumption |
| Infected, ICU | 0.025 | Assumption |
| Recovered | 9.425 | 11 |
| **Average time spent in state (days)** |  |  |
| Exposed | 3 | 12 |
| Asymptomatic ** | 6 | 12, 13 |
| Mild-Moderate | 6 | 12, 14 |
| Severe | 4 | 12, 14 |
| Critical | 14 | 12, 14 |
| **Likelihood of state-to-state progression (%)** |  |  |
| Exposed to Asymptomatic | 100 | Assumption |
| Asymptomatic to Mild/Moderate | 60 | 12 |
| Mild-Moderate to Severe | 10 | Assumption |
| Severe to Critical | 5 | Assumption |
| **Recovery (likelihood of recovering from state, %)** |  |  |
| Asymptomatic | 40 | Assumption |
| Mild-Moderate | 89 | Assumption |
| Severe | 90 | Assumption |
| Critical | 85 | Assumption |
| **Mortality (likelihood of death from state, %)** |  |  |
| Asymptomatic | 0 |  |
| Mild-Moderate | 1 | 15, 16 |
| Severe | 5 | Assumption |
| Critical | 15 | Assumption |
| **Time horizon (days)** | 60 | Assumption |
| **Population epidemiology** |  |  |
| Effective reproduction number, $R_t$ | 1.3 (0.9 – 2.1) | Assumption |
| **Test characteristics** | **Base case (range)** |  |
| Testing frequency (days) | 7 (1 – 15) | Assumption |
| Sensitivity (%) | 80 (70-95) | Assumption |
| Specificity (%) | 95 (90-99) | Assumption |
| Time to confirmatory test result return (days) | 3 | Assumption |
| **Costs (\$)** | **Base case (range)** | |
| Basic test kit | 5 (1 - 10) | Assumption |
| Confirmatory test kit | 20 (5 – 50) | Assumption |
| Cost of hospitalization | \$1,000/day | Assumption |
| Cost of ICU stay | \$2,500/day | Assumption |
| Productivity loss | \$180/day spent in isolation | 17, 18 |
| Value per statistical life (VSL) | \$5.3 - \$17.3 million | 10 |

